# Neonatal transport practices and effectiveness of the use of low-cost interventions on the morbidity and mortality of transported neonates in Sub-Saharan Africa: A systematic review protocol

**DOI:** 10.1101/2022.08.24.22279167

**Authors:** Emmanuel Okai, Hora Soltani, Frankie J Fair

## Abstract

**Objectives:** Poor pre-transport and intra-transport care are associated with early neonatal morbidity and mortality. The objective of this study is to produce a critical overview of research on neonatal transport practices, the use of any low-cost interventions, including Kangaroo mother care (KMC) employed in neonatal transport on the outcomes of neonates transported for specialized care in Sub-Saharan Africa, through a systematic review and narrative synthesis.

**Study design:** The study design is a protocol for systematic review

**Methods:** A comprehensive literature search will be conducted using the electronic databases of CINAHL, EMBASE, MEDLINE, Web of Science, as well as Google Scholar. A manual search will be carried out of the reference lists of eligible studies for relevant papers. The search strategy will include combining three key blocks of terms, namely: ‘neonates’, ‘transport’ and ‘Sub Saharan Africa,’ using database-specific subject headings and text words. Two independent reviewers will screen and assess data quality and extraction for synthesis. Any disagreements during the data extraction and quality assessment stages will be determined by consensus with the involvement of a third reviewer. Study quality will be assessed using the Mixed-Methods Appraisal Tool (MMAT); a narrative synthesis will be undertaken to integrate and summarise findings. Subgroup analysis will be done where data is available within countries with different groups of neonates and modes of transportation.

**Ethics and dissemination:** Approval by a research ethics committee is not required for this study since the review only includes published and publicly accessible data. Findings from this review will inform research into the use of Kangaroo Mother Care (KMC) in neonatal transport in Ghana. We will publish our findings in a peer-reviewed journal as per Preferred Reporting for Systematic Review and Meta-analysis (PRISMA) 2020 Guidance

**Registration:** PROSPERO registration CRD42022352401

## Background

Globally, significant progress has been made in child survival since the institution of the millennium development goals (MDGs), over two decades ago to tackle the indignity of poverty. Progress in reducing newborn mortality has however lagged behind the reduction in overall deaths in children under the age of 5 years.

The global under-5 mortality rate (U5MR) fell from 90·6 deaths per 1000 live births in 1990 to 42·5 per 1000 live births in 2015. During the same period, the annual number of under-5 deaths dropped from 12·7 million to 5·9 million (1), significantly below the 2015 (MDG) target of a reduction by two-thirds. Comparatively the decline in neonatal mortality has been slower than that of post-neonatal under-five mortality: 47 per cent compared with 58 per cent globally. The portion of neonatal deaths among overall under-five deaths increased from 40% in 1990 to 45% in 2015 (2) with several regions in the worse hit areas exceeding 50 per cent mortality (3). The neonatal mortality rate fell from 36 deaths per 1,000 live births in 1990 to 19 in 2015 and the number of neonatal deaths declined from 5.1 million in 1990 to 2.7 million in 2015 (2).

The Sustainable Development Goals (SDGs) have a child survival target for all countries to achieve a U5MR of 25 or fewer deaths per 1000 livebirths by 2030, (SDG target 3.2.1) and a neonatal mortality rate (NMR) of at least 12 per1000 livebirths by 2030 (SDG target 3.2.2) (4). Low-middle income countries (LMICs) bear the major brunt of this health disparity with Sub-Saharan Africa leading with a neonatal mortality rate of 27 deaths per 1000 live births accounting for 43% of global new-born deaths, followed by central and southern Asia (23 deaths per 1000 livebirths) (5)

The primary causes of neonatal mortality (prematurity, congenital anomalies, birth asphyxia, and severe infections) have received significant attention in terms of efforts toward their prevention but these must be complemented by the development of robust systems to care for sick new-borns, including safe and appropriate transfer processes.

### Neonatal Health service delivery and referral systems in LMICs

LMICs have significant challenges in almost all of the health system building blocks defined by the World Health Organization. These are service delivery, health workforce, health management information systems, access to essential medicines, financing, leadership and governance, and community engagement (6). Readily available access to skilled attendance at birth remains a major challenge in many resource-constrained settings (7) Although the pattern of health facility deliveries has generally increased in many countries, at least half of all births in developing countries occur in the absence of skilled birth attendants (8). The consequence of this disparity in health facility delivery translates into many neonates in resource-constrained settings being delivered in settings without adequate resources for newborn care, in suboptimal and often indeterminate conditions and would require transportation to better-equipped facilities where appropriate special or intensive care can be provided.

According to WHO statistics, about one-third of neonatal deaths in 2019 occurred on the day of birth with close to three quarters happening in the first 7-days (9). Many newborn deaths are a consequence of a lack of contact with the health system or a lack of transfer systems to improve access to higher-level facilities (10) (11) (12). Prompt attention to and referral of sick newborns might significantly contribute to a reduction in the high mortality occurring on the day of birth (13).

### Neonatal Transport in Lower-Middle income countries (LMICs)

Poor pre-transport and intra-transport care are associated with early neonatal morbidity and mortality. Serious neonatal morbidity particularly hypothermia, hypoglycaemia, hypoxia, and hyperthermia may result from poor transportation and inadequate care resulting in an increased risk of neonatal mortality (14). Undoubtedly, inutero transfer is the safest and most secure form of transfer, but the unpredictability of preterm delivery and perinatal illness has resulted in the continued need for an effective neonatal transport system (15) Sick newborns within the catchment areas of established specialized neonatal centres, remain at risk of adverse outcomes because of poor access, contributed to in part, by lack of coordinated neonatal transport service and oftentimes unsafe transfer practices. A structured and organized neonatal transport system can improve outcomes for newborns at high risk of adverse conditions who are delivered far from the established tertiary centres (16).

The Sub-Saharan sub-region has differing statistics, economically and in health compared to other regions considered as low-middle income. 40% of the world’s population currently live below the US$1.90-a-day poverty line with Sub-Saharan Africa accounting for 66 per cent of the global extreme poor population (17).

Challenges in every sector of the health system regarding newborn care and the absence of well-established ambulance service in most LMICs lend itself to the need to explore the possibility of employing cost-effective but evidence-based, life-saving processes in the neonatal transfer process which would inform technical guidelines for use by health care professionals, facilities, and organizations responsible for setting health system priorities and policies surrounding the transport of neonates in LMICs. The use of Kangaroo mother care and other alternative methods of thermal stability for neonatal transport is not well studied in LMICs, yet, this has been one of the areas proposed for future research in several published studies on neonatal transport

### Objectives

To systematically review the literature on neonatal transport practices in Sub-Saharan Africa and its contribution to morbidity and mortality in neonates transported for specialized care.

Review questions

1. What transport organizations and practices are available for neonatal transport into hospitals in Sub-Saharan Africa?
2. What evidence-based, low-cost interventions including Kangaroo mother care (KMC) are employed in neonatal transport in the region and how do these impact on morbidity and mortality of transported neonates?

## METHODS

### Study design

The study design is a protocol for a systematic review. It will include peer-reviewed primary empirical quantitative, qualitative, and mixed methods studies. The study protocol for this systematic review is prepared according to the guidelines given by the Preferred Reporting Items for Systematic Review and Meta-Analysis Protocols (PRISMA-P) statement.

### Participants or population

Any study primarily focused on all neonates, defined as newborn infants with a chronological age of 28 days or less without regard to the postmenstrual age at birth, who are referred from the community or a primary health care facility to a designated facility where specialized neonatal care is available irrespective of the distance

### Intervention(s), exposure(s)

All interventions to improve the outcome of referred neonates are relevant to this review. These include all pre-transport interventions e.g., training in the communication of medical personnel, inter-facility pre-referral communication, interventions that aim to refine or upgrade already existing methods of transport, and the introduction of new methods or equipment for transport and methods to improve physiologic stability.

### Comparator(s)/control

We will include all articles, whether or not a control group is used.

### Context

Articles from all 49 Sub-Saharan African countries studying the transfer of neonates from primary, secondary or tertiary settings to another healthcare facility for continuing care purposes.

Any type of transport will be considered: ground, air, repatriation, wait and return appointment.

### Main outcome(s)

The primary objective of this review is to synthesize evidence on the outcomes (morbidity and mortality) of the various transport practices of neonates that require inter-facility or community-to-hospital transport in Sub-Saharan Africa

### Additional outcome(s)

A secondary objective is to identify any evidence-based cost-effective intervention employed in the transport of neonates in the region, and define components of the interventions, such as at what stage they are employed (pre-transfer, during, or post-transfer), by whom and what outcomes were measured.

### Inclusion criteria

All peer-reviewed publications on neonatal transport, describing transport modalities/processes and outcomes within sub-Saharan Africa will be assessed. There will be no time restrictions for publications; no limit to language, however, searches will be restricted to only studies from Sub-Saharan Africa.

### Exclusion Criteria

Case reports, protocols, editorials, commentaries, or studies on neonatal transport not published from Sub-Saharan Africa. For studies with uncertainty about meeting inclusion criteria, or where relevant data cannot be extracted from the study report, the authors will be contacted for additional information and/or further data. If this is not forthcoming the study will be excluded. In the case of studies of mixed samples where only a proportion of the study population meets the inclusion criteria (e.g., only a proportion of the transported infants are neonates, or communication was partly about the maternal pre-labour transfer), the study will be included if relevant data can be extracted. The authors will be contacted if relevant data cannot be extracted, if this is not available, then the study will be excluded.

### Settings

For this review, studies from all 49 countries which are geographically in the subregion, including Djibouti which is not included in the world bank repository only because of administrative reasons (18), will be considered.

### Search strategy

A comprehensive search of the following databases will be undertaken for the identification of peer-reviewed literature: CINAHL, EMBASE, MEDLINE, and Web of Science, as well as Google Scholar and reference lists of included articles.

The search strategy will include terms around transport or transfer, neonate, and interventions. Boolean logic will be applied to ensure a comprehensive combination of search terms.

The searches will be conducted between 11^th^ and 22^nd^ April 2022 and a re-run before the final analysis conducted between 15^th^ and 26^th^ August 2022. No restrictions will be placed on the publication date or language of publication and searches will be conducted for relevant papers published up to August 2022.

However, the studies included will be restricted to only those from Sub-Saharan Africa.

### Data extraction (selection and coding)

Articles retrieved using the search strategy will initially be screened by title and abstract against the inclusion criteria. Full texts will then be retrieved and assessed for eligibility. The screening will be undertaken by the lead reviewer (EO) and checked by two other reviewers with any disagreement over potential article relevance resolved through discussion.

Study authors will be contacted via email correspondence for any relevant missing data or clarification. Duplicates will be eliminated using RefWorks citation manager.

Once eligible papers have been identified, they will be read in full text independently by two reviewers for inclusion and the primary reviewer (EO) will extract study characteristics and outcome data from the included studies using a predefined data extraction form (Excel spreadsheet) which will be subsequently checked for accuracy by two other reviewers.

Reasons for exclusion of studies will be provided and screening of search results will be presented in a PRISMA flow chart.

Data would be extracted under the following headings

Title, author, year of publication, country of origin, study design, Population (size, term, preterm, low birth weight). Other specific data to be extracted include,

Type of transfer (ambulance-ground/air, public transport, walking, cycle); Accompanying parents or health personnel.

Intervention types (e.g., KMC, focussed on communication, physical/practical, social, economic, psychological or emotional wellbeing);

Intervention timing (pre-transfer, during transfer, post-transfer short or long-term); Intervention by whom (transport team member, midwife/neonatal unit staff, specialised staff such as a psychologist, occupational therapist, social worker, etc.);

Intervention effectiveness testing (what scale/tool used and effectiveness rating), Outcomes: Mortality, hypothermia, hypoglycaemia, hypoxia, apnoea, respiratory distress

### Risk of bias assessment

As the review will include quantitative, qualitative, and mixed methods studies, the quality of included studies will be assessed independently by two reviewers using the Mixed-Methods Appraisal Tool (MMAT), V.2018 (19). The MMAT includes two screening questions, followed by five categories of study design assessment including qualitative, quantitative – non-randomised, randomised, descriptive, and mixed methods. Within each study category, there are five methodological quality criteria. These criteria relate to the appropriateness of methodology, data collection techniques and data analysis techniques. Each included study will be appraised thoroughly using the entire assessment criteria. independently by two reviewers, with any disagreements discussed with a third reviewer. No studies will be excluded based on their quality, but the narrative synthesis will comment on the quality of the identified studies.

### Strategy for data synthesis

Due to the expected heterogeneity of studies, a narrative synthesis of the results will be performed. This will include a narrative discussion of methods of neonatal transportation and the adequacy or otherwise of the transfer process and the outcomes for the transported neonates.

Additionally, cost-effective interventions employed, their adaption or otherwise and effectiveness in the transport process would be described.

Where sufficient and relevant data is available, subgroup analysis would be undertaken for the various categories of neonates (term, preterm and low birth weight) in the context of the varying transport modalities

The screening results will be reported in a PRISMA flow diagram.

## Conclusion

This systematic review of neonatal transport in Sub-Saharan Africa will provide a detailed summary of current neonatal transport modalities and organization as well as evidence for the effectiveness of low-cost interventions employed to improve the outcomes of transported neonates from primary health care into specialized care centres.

## Data Availability

N/A

## Acknowledgements

Prof. Dilly Anumba. Department of Oncology and Metabolism, University of Sheffield, UK Prof. Eugene Kofuor Maafo Darteh, Department of Population and Health, University of Cape Coast, Cape Coast, Ghana

## Subject index terms

Infant; Infant, Newborn; Intensive Care Units, Neonatal; Intensive Care, Neonatal; Transfer, Transport Sub-Saharan Africa, developing countries, LMICs

## Author statements

### Ethical approval

Not required for protocol papers of systematic review

